# Trends and Heterogeneity in Having a Usual Source of Care Among U.S. Children: Evidence from the Medical Expenditure Panel Survey, 2015-2024

**DOI:** 10.64898/2026.09.15.26363170

**Authors:** Bing Han, Bowen Feng, Chuntao Zhao, Richard S. Han, Jingyun Yang

**Author notes:** Corresponding author Bing Han, Department of Research and Evaluation, Kaiser Permanente Southern California, 100 S Los Robles Ave., Pasadena, CA 91101, U.S.A.

## Abstract

**Background:** Having a usual source of care (USC) supports continuity and access to health services. Recent national trends among U.S. children have not been comprehensively characterized. We examined changes in USC prevalence, subgroup heterogeneity, and reported reasons for not having a USC using the 2015-2024 Medical Expenditure Panel Survey.

**Methods:** We analyzed the 2015-2024 Medical Expenditure Panel Survey Household Full-Year Consolidated Data. Annual prevalence rates were estimated for children aged 0 to 17 years overall and by sex, age group, race and ethnicity, and insurance status. The reported reasons for not having a USC were grouped to low perceived need, preference or self-management, access or continuity barrier, and other reasons. The distribution of grouped reasons was analyzed for each study year. Temporal differences in yearly estimates were evaluated using two-sample z-tests between 2015 and 2019 and between 2015 and 2024, and average annual changes were estimated using generalized linear regression.

**Results:** Overall USC prevalence declined from 92.9% (95% confidence interval [CI], 91.9%, 93.8%) in 2015 to 87.2% (95% CI, 85.1%, 89.2%) in 2024. The estimated average annual change was −0.8 percentage points (95% CI, −1.0 to −0.6). Declines were observed across most subgroups and were largest among uninsured children, whose prevalence decreased from 78.4% to 53.6%. Low perceived need remained the most common reported reason for not having a USC, while the proportion reporting access or continuity barriers decreased and the proportion classified as other reasons increased.

**Conclusions:** Estimates based on the Medical Expenditure Panel Survey indicated a substantial decline in the prevalence of having a USC among U.S. children, with notable heterogeneity by insurance status and race and ethnicity. Further research is needed to determine whether the decline reflects changing access, organization and use of pediatric care, survey measurement, or a combination of these factors.

## INTRODUCTION

A usual source of care (USC) is a particular medical professional, doctor’s office, clinic, health center, or other place where a person usually goes when sick or in need of health advice, generally excluding hospital emergency department.^1^ For children, a USC provides a regular point of contact with the health care system and is an important indicator of consistent access to care.^2–5^ Having a USC supports continuity of care, a central feature of the pediatric medical-home model endorsed by the American Academy of Pediatrics.^6–9^ Prior empirical studies have operationalized a USC as a necessary condition to establish parent-reported or patient-reported medical home status.^9–19^

Earlier national estimates suggested that most U.S. children had a USC, although important disparities and heterogeneity remained. Using the Medical Expenditure Panel Survey (MEPS) data, Zewde and Berdahl reported that the percentage of children without a USC varied from 7.9% to 11.0% between 2004 and 2014, and was the lowest in 2013 and 2014 at 7.9%.^2^ Children who were uninsured or from lower-income families were more likely to lack a USC.^2^ Prior studies have also documented that children’s USC status differed by insurance coverage, sociodemographic groups, and parental USC status.^2,3,20^

The delivery of pediatric care has evolved substantially in recent years. Acute nonemergency care has been increasingly provided through urgent care, retail clinics, various telehealth platforms, and other settings.^6,7,21–25^ The COVID-19 pandemic introduced a sharp interruption to pediatric primary care visits, although preventive visits later rebounded.^26,27^

Recent official estimates based on MEPS indicated that 10.6% and 13.9% of the U.S. population aged 0-18 did not have a USC in 2020 and 2021, respectively, a notable increase from 2014.^28,29^ The heterogeneity and disparities in having a USC may have increased in recent years. However, rigorous quantitative evidence on the national trends and subpopulation differences remains lacking.

Using the nationally representative MEPS data from 2015 through 2024, we examined national trends in the percentage of having a USC among U.S. children aged 0-17 and by subpopulations defined by age group, sex, insurance status, and race/ethnicity. We also examined trends in parent-reported reasons for lacking a USC. This study aimed to provide descriptive statistical evidence and was not intended for identifying the causes of observed trends.

## METHODS

### Data source and study sample

MEPS is a nationally representative survey for the U.S. civilian noninstitutionalized population and conducted by the Agency for Healthcare Research and Quality. We used the MEPS Household Full-Year Consolidated Data Files from 2015 through 2024. Each annual file is a nationally representative cross-sectional sample for that calendar year and generally includes two partially overlapping two-year panels: one that began in the prior year and one that began in the current year. All data files are publicly available on the MEPS official website https://meps.ahrq.gov/mepsweb/index.jsp. For each calendar year in the study period, the analytic sample included children younger than 18 years as of December 31 who were eligible for the MEPS Access to Care section and provided a non-missing response to the USC survey question.

### Study variables

The primary variable of interest was the respondent-reported status of whether a child had a USC in the survey calendar year. In accordance with the literature, those who reported their USC’s practice location was the emergency department (ER) in a hospital were reclassified as not having a USC.^2,11,13^ Child-level characteristics for subgrouping included integer-valued age at the end of the year and stratified to three age categories (0-4, 5-12, 13-17 years), sex (male, female), race/ethnicity (Hispanic of all races, non-Hispanic single-race White, non-Hispanic single-race African American, and other races/ethnicities, shortened as Hispanic, White, Black, and Others, hereafter). and insurance status (private, public, uninsured).

For those without a USC, we further examined the reported reasons for not having one. The original categorical survey responses in MEPS were consolidated into four broad groups: low perceived need (i.e., “seldom or never sick” in original response), preference or self-management (e.g., “don’t use doctors/treat self” in original response), access or continuity barrier (e.g., “cost of medical care” in original response), and other reasons that cannot be classified into the three preceding groups. Missing responses were grouped into other reasons. Details of the grouping were included in the supplementary materials (Supplement Table 1).

All study variables, as well as survey design variables (strata, cluster, and sampling weights), were available in MEPS throughout the study period 2015-2024. The MEPS survey underwent a major instrument redesign and coding restructure in 2018. The coding and response format of the core USC indicator, child-level characteristics, and survey design variables remained consistent before and after the redesign. The coding of USC practice location was revamped in the MEPS instrument redesign. However, the ER coding did not have major changes. Reporting ER in a hospital as the location of a USC was always very rare before and after the MEPS redesign. The reasons for not having a USC were also changed in the MEPS instrument redesign. We therefore tried to define the four broad groups to facilitate comparisons over time. Details of the MEPS variable name, coding, and grouping were in the supplementary materials (Supplement table 1).

### Statistical analysis

In each calendar year, we estimated the overall prevalence of having a USC and subgroup prevalence by sex, age strata (<5, 5-12, and 13-17 years), race/ethnicity, and health insurance status. Among children without a USC, we also estimated the annual distribution of the four grouped reasons for not having a USC. All point estimates and 95% confidence intervals (CI) accounted for MEPS complex survey designs (stratification, clustering, and survey weights).

Potential changes over time were first compared by two-sample z-tests between 2015 and 2019 and between 2015 and 2024. We also evaluated crude linear temporal trends using inverse-variance weighted generalized linear regression of the annual survey-weighted prevalence estimates on calendar year, with weights based on the inverse of the squared design-based standard errors. The estimated annual slope was intended as a descriptive summary of the 10-year period and was not interpreted as evidence that the underlying temporal relationship was strictly linear.. All statistical estimates were exploratory and descriptive. Therefore, we did not adjust for simultaneous inference. 95% CI not covering the null value was considered as statistically significant.

Sensitivity analysis included a relaxed definition of USC without restricting to non-ER locations. We also fitted the weighted linear regression with an autoregressive AR(1) correlation structure to test the potential serial correlations among annual MEPS data.

## RESULTS

The yearly raw sample size of the study sample decreased steadily from 9,260 in 2015 to 3,389 in 2024 (Supplement table 2). Figure 1 presents the estimated overall and subgroup prevalence of having a USC among U.S. Children from 2015 through 2024 based on MEPS data. Overall prevalence declined from 92.9% (95% CI: 91.9%, 93.8%) in 2015 to 89.2% (95% CI: 87.6%, 90.8%) in 2019 and 87.2% (95% CI: 85.1%, 89.2%) in 2024. Overall prevalence of USC changed significantly by −3.6 percentage points (95% CI: −5.5, −1.8) from 2015 to 2019, and by −5.7 percentage points (95% CI: −7.9, −3.4) from 2015 to 2024 (Table 1). The average decline was −0.8 percentage points per year (95% CI: −1.0, −0.6) (Table 1).

**Figure 1.**
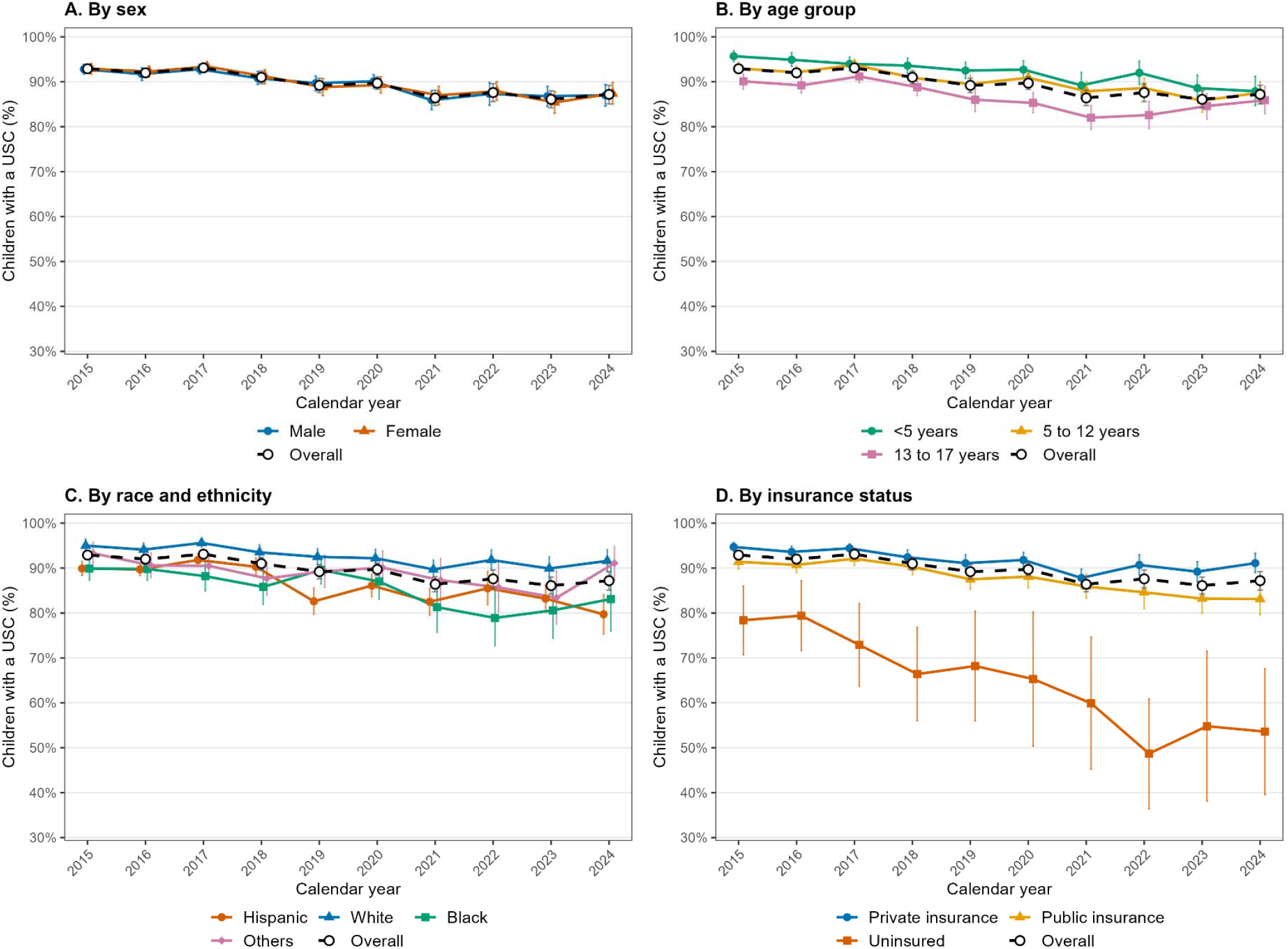
Annual estimates and 95% confidence intervals for prevalence of having a usual source of care among U.S. children based on Medical Expenditure Panel Survey 2015-2024.

**Table 1.** Estimated prevalence of having a usual source of care among U.S. children in 2015, 2019, and 2024 and changes over time: percentage points (95% CI)

|  | 2015 | 2019 | Difference<br>2019 vs. 2015 | 2024 | Difference<br>2024 vs. 2015 | Average<br>annual change |
| --- | --- | --- | --- | --- | --- | --- |
| Overall | 92.9<br>(91.9, 93.8) | 89.2<br>(87.6, 90.8) | -3.6<br>(-5.5, -1.8) | 87.2<br>(85.1, 89.2) | -5.7<br>(-7.9, -3.4) | -0.8<br>(-1.0, -0.6) |
| Male | 92.8<br>(91.7, 93.9) | 89.7<br>(88.0, 91.3) | -3.2<br>(-5.2, -1.2) | 87.0<br>(84.6, 89.4) | -5.9<br>(-8.5, -3.2) | -0.8<br>(-1.0, -0.5) |
| Female | 92.9<br>(91.7, 94.1) | 88.8<br>(86.9, 90.7) | -4.1<br>(-6.3, -1.9) | 87.4<br>(85.0, 89.8) | -5.5<br>(-8.2, -2.8) | -0.9<br>(-1.1, -0.6) |
| <5 years | 95.7<br>(94.5, 96.9) | 92.5<br>(90.7, 94.4) | -3.2<br>(-5.4, -1.0) | 87.9<br>(84.7, 91.2) | -7.8<br>(-11.3, -4.3) | -0.8<br>(-1.1, -0.5) |
| 5 to 12 years | 93.0<br>(91.8, 94.3) | 89.5<br>(87.6, 91.4) | -3.5<br>(-5.8, -1.3) | 87.6<br>(85.2, 90.0) | -5.4<br>(-8.1, -2.7) | -0.8<br>(-1.0, -0.5) |
| 13 to 17 years | 90.1<br>(88.4, 91.7) | 86.0<br>(83.4, 88.6) | -4.1<br>(-7.2, -1.1) | 85.9<br>(82.9, 89.0) | -4.2<br>(-7.6, -0.7) | -0.9<br>(-1.2, -0.6) |
| Hispanics | 89.9<br>(88.3, 91.5) | 82.6<br>(79.7, 85.6) | -7.3<br>(-10.6, -4.0) | 79.7<br>(75.3, 84.1) | -10.2<br>(-14.9, -5.5) | -1.1<br>(-1.4, -0.7) |
| White | 95.0<br>(93.6, 96.5) | 92.5<br>(90.7, 94.3) | -2.6<br>(-4.9, -0.2) | 91.6<br>(89.2, 94.1) | -3.4<br>(-6.3, -0.5) | -0.6<br>(-0.9, -0.4) |
| Black | 89.9<br>(87.3, 92.5) | 89.6<br>(86.5, 92.7) | -0.3<br>(-4.3, 3.8) | 83.1<br>(76.0, 90.1) | -6.8<br>(-14.3, 0.7) | -1.0<br>(-1.7, -0.4) |
| Others | 93.3<br>(90.9, 95.8) | 89.3<br>(85.7, 92.9) | -4.0<br>(-8.3, 0.3) | 91.1<br>(87.3, 94.8) | -2.3<br>(-6.8, 2.2) | -0.5<br>(-1.0, 0.0) |
| Private insurance | 94.7<br>(93.7, 95.7) | 91.1<br>(89.1, 93.0) | -3.6<br>(-5.8, -1.4) | 91.1<br>(89.0, 93.3) | -3.5<br>(-5.9, -1.2) | -0.6<br>(-0.9, -0.4) |
| Public insurance | 91.4<br>(89.8, 92.9) | 87.5<br>(85.3, 89.6) | -3.9<br>(-6.5, -1.2) | 83.1<br>(79.6, 86.7) | -8.2<br>(-12.1, -4.3) | -1.0<br>(-1.4, -0.7) |
| Uninsured | 78.4<br>(70.7, 86.0) | 68.2<br>(56.0, 80.4) | -10.2<br>(-24.6, 4.3) | 53.6<br>(39.6, 67.6) | -24.8<br>(-40.7, -8.8) | -3.4<br>(-5.0, -1.8) |

Subgroup analysis showed mostly consistent cross-sectional patterns in all years. There was no notable cross-sectional difference in USC prevalence by sex in any given year. In 2024, prevalence was 87.0% (95% CI: 84.6%, 89.4%) among boys and 87.4% (95% CI: 85.0%, 89.8%) among girls. Prevalence was generally lower with children’s age. In 2024, it was 87.9% (95% CI: 84.7%, 91.2%) for children aged 0-4 years, 87.6% (95% CI: 85.2%, 90.0%) for those aged 5-12 years, and 85.9% (95% CI: 82.9%, 89.0%) for those aged 13-17 years (Table 1). Compared with White children, prevalence was lower in Hispanic and Black children. The 2024 estimates (Table 1) were 91.6% for White (95% CI: 89.2%, 94.1%), 79.7% for Hispanic (75.3%, 84.1%), 83.1% for Black (76.0%, 90.1%), and 91.1% for Others (87.3%, 94.8%). Prevalence was the highest for privately insured children, publicly insured had notably lower prevalence rates than privately insured, and the uninsured subgroup had substantially lower prevalence of USC. In 2024, prevalence was 91.1% for privately insured (95% CI: 89.0%, 93.3%), 83.1% for publicly insured (95% CI: 79.6%, 86.7%), and 53.6% for uninsured (95% CI: 39.6%, 67.6%) (Table 1).

Prevalence of USC had generally declined throughout the study period in most subgroups with an extent similar to the pattern of overall prevalence. The uninsured children had the most pronounced decline over time from 78.4% in 2015 to 53.6% in 2024, with an average decline of 3.4 percentage points per year (95% CI: 1.8, 5.0). However, the CIs for uninsured children were generally wide due to their small sample size (Figure 1). In contrast, publicly insured declined by 1.0 percentage points per year (95% CI: 0.7, 1.4) and privately insured declined by 0.6 percentage points per year (95% CI: 0.4, 0.9). Estimated annual declines were numerically larger among Hispanic and Black children, at 1.1 and 1.0 percentage points per year, respectively, than among White children and Others, for whom the estimated decline was 0.6 percentage points per year in both groups.

Figure 2 shows the conditional distribution of grouped reasons for not having a USC among children without a USC. Low perceived need was always the most common reason throughout the study period. Its estimated proportion increased from 47.9% in 2015 (95% CI: 41.5%, 54.3%) to 55.4% in 2019 (95% CI: 49.0%, 61.7%) was similar in 2024 (54.1%, 95% CI: 46.9%, 61.3%), with a significant average increase of 1.1 percentage points per year (95% CI: 0.1, 2.1) (Table 2).

**Figure 2.**
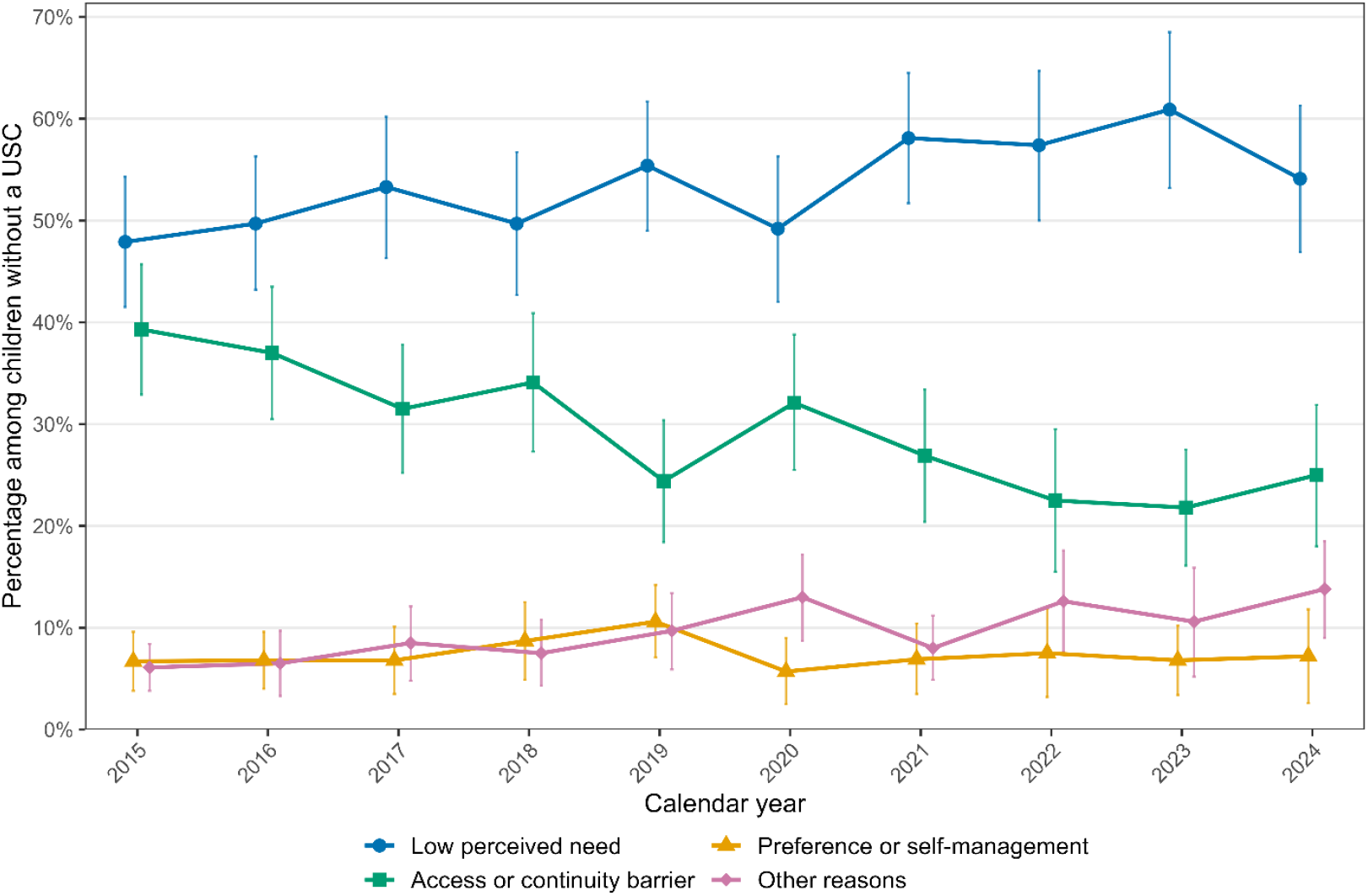
Annual estimates and 95% confidence intervals for grouped reasons among U.S. children not having a usual source of care based on Medical Expenditure Panel Survey 2015-2024.

**Table 2.**
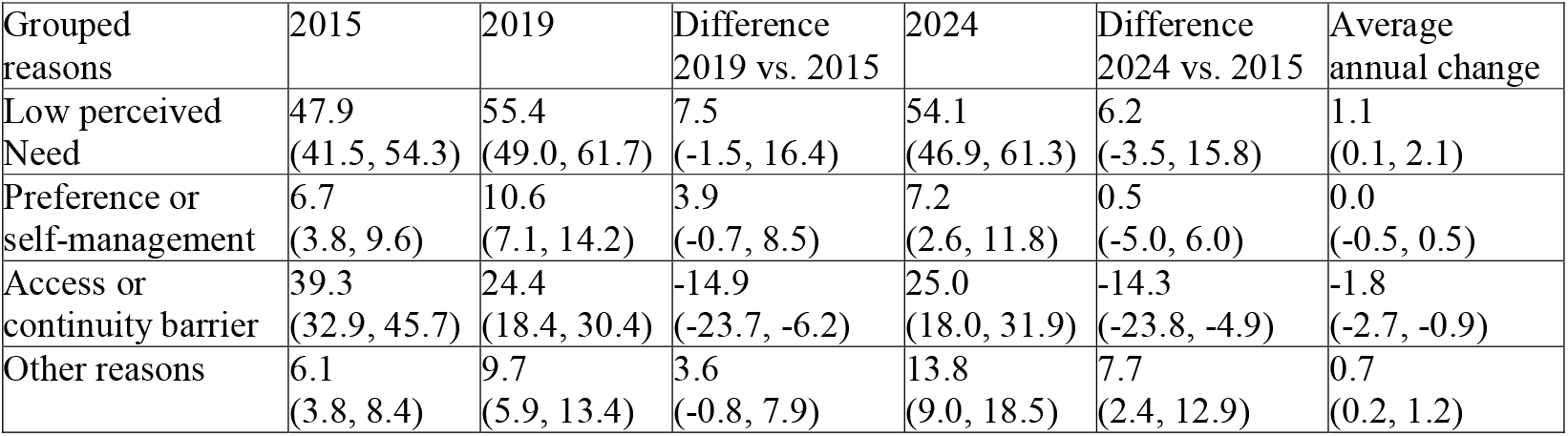
Estimated distribution of reported reasons for not having a usual source of care (USC) among U.S. children without a USC in 2015, 2019, and 2024 and changes over time: percentage points (95% CI).

| Grouped reasons | 2015 | 2019 | Difference 2019 vs. 2015 | 2024 | Difference 2024 vs. 2015 | Average annual change |
| --- | --- | --- | --- | --- | --- | --- |
| Low perceived Need | 47.9<br>(41.5, 54.3) | 55.4<br>(49.0, 61.7) | 7.5<br>(-1.5, 16.4) | 54.1<br>(46.9, 61.3) | 6.2<br>(-3.5, 15.8) | 1.1<br>(0.1, 2.1) |
| Preference or self-management | 6.7<br>(3.8, 9.6) | 10.6<br>(7.1, 14.2) | 3.9<br>(-0.7, 8.5) | 7.2<br>(2.6, 11.8) | 0.5<br>(-5.0, 6.0) | 0.0<br>(-0.5, 0.5) |
| Access or continuity barrier | 39.3<br>(32.9, 45.7) | 24.4<br>(18.4, 30.4) | -14.9<br>(-23.7, -6.2) | 25.0<br>(18.0, 31.9) | -14.3<br>(-23.8, -4.9) | -1.8<br>(-2.7, -0.9) |
| Other reasons | 6.1<br>(3.8, 8.4) | 9.7<br>(5.9, 13.4) | 3.6<br>(-0.8, 7.9) | 13.8<br>(9.0, 18.5) | 7.7<br>(2.4, 12.9) | 0.7<br>(0.2, 1.2) |

Access or continuity barriers were always the second most common reason, whose estimated proportion, however, declined from 39.3% in 2015 (95% CI: 32.9%, 45.7%) to 24.4% in 2019 (95% CI: 18.4%, 30.4%) and increased slightly to 25.0% in 2024 (95% CI: 18.0%, 31.9%). The estimated average annual change was significant (−1.8 percentage points per year; 95% CI: −2.7, −0.9) (Table 2).

Figure 2 and Table 2 also show that preference or self-management was uncommon, ranging from 5% to 11% with no notable linear change over time. The grouped all other reasons were also uncommon. However, its estimated proportion increased from 6.1% in 2015 (95% CI: 3.8%, 8.4%) to 9.7% in 2019 (95% CI: 5.9%, 13.4%) and to 13.8% in 2024 (95% CI: 9.0%, 18.5%), where the average annual increase estimate was also significant (0.7 percentage points per year; 95% CI: 0.2, 1.2) (Table 2).

Detailed yearly estimates and 95% CI were reported in supplementary materials (Supplement tables 3 and 4). Sensitivity analyses using alternative USC definitions and correlation assumptions over years yielded only minor numerical differences in point estimates and did not alter statistical inference or interpretation of the findings. Therefore, detailed sensitivity analysis results are omitted.

## DISCUSSION

In this nationally representative analysis of U.S. children, we found that the prevalence of having a USC declined significantly from 2015 through 2024. Yearly estimates series did not show a clear, isolated, and one-year discontinuity followed by stabilization in 2018 when the MEPS instrument was restructured. We also did not find a sudden and rapid change after the onset of the COVID-19 pandemic, but the year-to-year variation after 2020 may be slightly larger. These findings extend earlier MEPS evidence showing that 7.9% to 11.0% of children lacked a USC between 2004 and 2014 and show a concerning signal.^2^ The agreement of our results with published MEPS-based MACStats estimates also supports the reproducibility of the magnitude observed in the later years.^28,29^

The decline in USC prevalence during the study period occurred across most demographic and insurance subgroups, though the magnitude varied. The uninsured group experienced the steepest decline. The estimated declines were also larger for Hispanic and Black children than for White children.

Cross-sectionally, older, Hispanic, Black, and uninsured children had lower prevalence of USC than their counterparts, correspondingly. These patterns are consistent with prior evidence that insurance coverage, family resources, race and ethnicity, and parental access to a USC are associated with children’s access to a regular source of care.^2,3,20,28,29^

The declining USC prevalence was accompanied by an increase in the proportion of low perceived need, a decrease in the proportion of access or continuity barriers, and an increase in the proportion of other reasons. Note that the estimated reason distribution was conditioning on children not having a USC. A decreasing or stable proportion for a grouped reason coexisted with the expansion of children without a USC. In addition, the broad reason categories may not capture emerging forms of fragmented or episodic care, including reliance on urgent care, retail clinics, telehealth platforms, or multiple sites that families may not describe as barriers.^6,7,21–25^

It is worth noting that our study findings based on MEPS data differ systematically from recent estimates based on the National Health Interview Survey (NHIS), which had substantially higher USC prevalence estimates.^30^ While it is not uncommon for national surveys from different agencies to have systematic differences,^28,29,31,32^ the difference between MEPS and NHIS in Children’s USC status has been well documented and may reflect differences in the questionnaire wording and survey methods. MEPS always asks whether “there is a particular doctor’s office, clinic, health center, or other place the child usually goes when sick or needing health advice”, whereas the recently redesigned NHIS asks whether “there is a place the child usually goes when sick and needing health care”, besides technical differences in sampling design, context, administration, and other aspects. Our estimates should therefore be interpreted as evidence of a decline within the MEPS framework and should not be generalized to other contexts without due caution. Future methodological studies are still needed for the systematic differences between these two important national surveys.

This study has a few important internal limitations. First, the study was descriptive and did not intend to discuss any causal mechanisms for temporal changes or cross-sectional heterogeneity. Changes in health care delivery, health insurance policy, and social and physiological preferences, as well as the COVID-19 pandemic may all be relevant, but our descriptive analyses were not able to investigate their contributions. Second, the 2018 MEPS redesign remains a weakness. Despite the sensitivity analysis, we could not fully eliminate the potential impact of survey instrument redesign. Third, the broad reason groups may not capture emerging forms of fragmented or episodic care, including reliance on urgent care, retail clinics, telehealth platforms, or multiple sites that families may not describe as barriers.^6,7,21–25^ Fourth, some subgroup estimates, especially among uninsured and racial/ethnic minority children, were imprecise, as reflected by the wide CIs in these subgroups. Fifth, the linear temporal slope was a crude and naïve summary of the 10-year study period, which should not be interpreted as a strict linear relationship and should not be extrapolated in time. Lastly, all survey variables were subject to various reporting biases and errors, and the small sample size in some subgroups reduced the precision in statistical inference.

In summary, despite the limitations, this study identified a broad weakening of attachment to a regular source of care and substantial heterogeneity across population groups based on the MEPS framework. Future studies are needed to disentangle and ascertain the mechanisms underlying the decline and causal barriers that explain subgroup heterogeneity and disparities.

## Data Availability

All raw data are available at https://meps.ahrq.gov/mepsweb/index.jsp. All numerical result data are contained in the paper and supplementary materials.

## DECLARATION

This paper is a secondary analysis of publicly available national survey data. Ethics approval and participant consent were not applicable. There is no funding source to disclose. The authors declare no conflict of interest.

