## Supplementary materials for "Trends and Heterogeneity in Having a Usual Source of Care Among U.S. Children: Evidence from the Medical Expenditure Panel Survey, 2015-2024"

**Supplementary Materials for** “**U.S. Children’s Access to A Usual Source of Care: 2015–2024”**

**Supplement table 1. MEPS variable names, recoding rules, and meanings.**

| Variable | MEPS variable name | Recoding rules and meanings |
| --- | --- | --- |
| Flag for having a USC | HAVEUS42 | 1: yes; 2: no; all other values: missing  Children with missing values were ineligible for the Access to Care section in MEPS and excluded from all analyses. |
| Age | AGE16X ~ AGE23X  Variable names change across survey years | Integer-valued age at year end (December 31). |
| Sex | SEX | 1: male; 2: female |
| Race / ethnicity | RACETHX |  |
| Insurance status | INSCOV16 ~ INSCOV23  Variable names change across survey years | During the year:  1: any private insurance;  2: public insurance only;  3: uninsured |
| Main reasons for not having USC (2018 and later) | YNOUSC42_M18 | Low perceived needs  1: Seldom or never sick  Preference or self-management  7: Likes to go different places for different needs  8: Don’t use doctors / treat self  Access or continuity barrier  2: Recently moved to area  3: Just changed insurance plans  4: No health insurance, other insurance-related issues  5: Don’t know where to go  6: USC in area no longer available  9: Cost of medical care  Other reasons  -8: Don’t know  -7: Refused  91: Other reasons |
| Main reasons for not having USC (before 2018) | YNOUSC42 | Low perceived needs  1: Seldom or never sick  Preference or self-management  6: Goes different places for diff needs  8: Don’t use Docs / treat self  14: Don’t like / Don’t true doctors  17: Self, relative, or friend is a doctor  20: Will not go to the doctor  23: Uses alternative care  Access or continuity barrier  2: Recently moved to area  3: Don’t know where to go  4: USC in area not available  5: Can’t find provider who speaks language  7: Just changed insurance plans  9: Cost of medical care  10: No health insurance  11: Job-related reasons  12: Looking for a new doctor / no doctor yet  13: USC doctor is somewhere else  21: Problems with time and transportation  24: Insurance-related reasons  Other reasons  -9: Not ascertained  -8: DK (Don’t know)  -7: Refused  15: Health-related reasons  16: Newborn – no doctor yet  19: Care available on job  22: Goes to hospital / emergency room / clinic  91: Other reasons |
| Stratum | VARSTR | MEPS stratum for variance estimation |
| Cluster | VARPSU | MEPS cluster from variance estimation |
| Sampling weight | PERWT15F ~ PERWT24F | Person-level survey weight |

**Supplement table 2. Unadjusted sample characteristics of the study sample pulled from the Medical Expenditure Panel Survey 2015:2024**

| N(%) | 2015 | 2016 | 2017 | 2018 | 2019 | 2020 | 2021 | 2022 | 2023 | 2024 |
| --- | --- | --- | --- | --- | --- | --- | --- | --- | --- | --- |
| Overall | 9,260 (100.0%) | 8,852 (100.0%) | 7,813 (100.0%) | 7,086 (100.0%) | 6,277 (100.0%) | 5,683 (100.0%) | 5,299 (100.0%) | 4,103 (100.0%) | 3,449 (100.0%) | 3,489 (100.0%) |
| Male | 4,793 (51.8%) | 4,484 (50.7%) | 3,973 (50.9%) | 3,656 (51.6%) | 3,234 (51.5%) | 2,926 (51.5%) | 2,741 (51.7%) | 2,141 (52.2%) | 1,757 (50.9%) | 1,814 (52.0%) |
| Female | 4,467 (48.2%) | 4,368 (49.3%) | 3,840 (49.1%) | 3,430 (48.4%) | 3,043 (48.5%) | 2,757 (48.5%) | 2,558 (48.3%) | 1,962 (47.8%) | 1,692 (49.1%) | 1,675 (48.0%) |
| <5 years | 2,193 (23.7%) | 2,088 (23.6%) | 1,825 (23.4%) | 1,668 (23.5%) | 1,459 (23.2%) | 1,271 (22.4%) | 1,150 (21.7%) | 868 (21.2%) | 748 (21.7%) | 774 (22.2%) |
| 5 to 12 years | 4,423 (47.8%) | 4,220 (47.7%) | 3,689 (47.2%) | 3,370 (47.6%) | 2,908 (46.3%) | 2,627 (46.2%) | 2,501 (47.2%) | 1,933 (47.1%) | 1,627 (47.2%) | 1,674 (48.0%) |
| 13 to 17 years | 2,644 (28.6%) | 2,544 (28.7%) | 2,299 (29.4%) | 2,048 (28.9%) | 1,910 (30.4%) | 1,785 (31.4%) | 1,648 (31.1%) | 1,302 (31.7%) | 1,074 (31.1%) | 1,041 (29.8%) |
| Hispanic | 3,918 (42.3%) | 3,720 (42.0%) | 2,972 (38.0%) | 2,339 (33.0%) | 2,029 (32.3%) | 1,942 (34.2%) | 1,860 (35.1%) | 1,247 (30.4%) | 1,032 (29.9%) | 1,042 (29.9%) |
| White | 2,430 (26.2%) | 2,435 (27.5%) | 2,660 (34.0%) | 2,914 (41.1%) | 2,643 (42.1%) | 2,279 (40.1%) | 2,114 (39.9%) | 1,726 (42.1%) | 1,486 (43.1%) | 1,561 (44.7%) |
| Black | 1,864 (20.1%) | 1,678 (19.0%) | 1,299 (16.6%) | 1,066 (15.0%) | 966 (15.4%) | 868 (15.3%) | 755 (14.2%) | 619 (15.1%) | 491 (14.2%) | 458 (13.1%) |
| Others | 1,048 (11.3%) | 1,019 (11.5%) | 882 (11.3%) | 767 (10.8%) | 639 (10.2%) | 594 (10.5%) | 570 (10.8%) | 511 (12.5%) | 440 (12.8%) | 428 (12.3%) |
| Private insurance | 3,635 (39.3%) | 3,522 (39.8%) | 3,492 (44.7%) | 3,569 (50.4%) | 3,237 (51.6%) | 2,785 (49.0%) | 2,598 (49.0%) | 2,159 (52.6%) | 1,820 (52.8%) | 1,877 (53.8%) |
| Public insurance | 5,274 (57.0%) | 5,000 (56.5%) | 4,121 (52.7%) | 3,317 (46.8%) | 2,882 (45.9%) | 2,730 (48.0%) | 2,576 (48.6%) | 1,842 (44.9%) | 1,533 (44.4%) | 1,487 (42.6%) |
| Uninsured | 351 (3.8%) | 330 (3.7%) | 200 (2.6%) | 220 (3.1%) | 158 (2.5%) | 168 (3.0%) | 125 (2.4%) | 102 (2.5%) | 96 (2.8%) | 125 (3.6%) |

**Supplement Table 3. Yearly prevalence rate estimates of U.S. children having a USC 2015-2024: cells are percentage (95% CI).**

|  | 2015 | 2016 | 2017 | 2018 | 2019 | 2020 | 2021 | 2022 | 2023 | 2024 |
| --- | --- | --- | --- | --- | --- | --- | --- | --- | --- | --- |
| Overall | 92.9  (91.9, 93.8) | 92.0  (91.0, 93.0) | 93.1  (92.2, 94.0) | 91.0 (89.6, 92.4) | 89.2 (87.6, 90.8) | 89.7 (88.3, 91.1) | 86.4 (84.7, 88.1) | 87.6 (85.6, 89.6) | 86.1 (84.2, 88.0) | 87.2 (85.1, 89.2) |
| Male | 92.8  (91.7, 93.9) | 91.7  (90.3, 93.1) | 92.8  (91.7, 93.9) | 90.8 (89.4, 92.3) | 89.7 (88.0, 91.3) | 90.1 (88.6, 91.6) | 85.9 (83.8, 88.0) | 87.3 (84.8, 89.8) | 86.8 (84.5, 89.0) | 87.0 (84.6, 89.4) |
| Female | 92.9  (91.7, 94.1) | 92.3  (91.1, 93.5) | 93.4  (92.3, 94.4) | 91.2 (89.6, 92.7) | 88.8 (86.9, 90.7) | 89.3 (87.5, 91.1) | 87.0 (84.9, 89.0) | 87.9 (85.9, 90.0) | 85.4 (83.0, 87.8) | 87.4 (85.0, 89.8) |
| <5 years | 95.7  (94.5, 96.9) | 94.9  (93.4, 96.5) | 94.0  (92.5, 95.5) | 93.6 (91.9, 95.3) | 92.5 (90.7, 94.4) | 92.7 (90.7, 94.7) | 89.2 (86.3, 92.1) | 92.0 (89.4, 94.6) | 88.6 (85.7, 91.5) | 87.9 (84.7, 91.2) |
| 5 to 12 years | 93.0  (91.8, 94.3) | 92.1  (90.8, 93.5) | 93.8  (92.7, 94.9) | 90.9 (89.3, 92.5) | 89.5 (87.6, 91.4) | 90.9 (89.2, 92.5) | 87.9 (85.8, 90.0) | 88.6 (86.3, 90.8) | 85.8 (83.2, 88.3) | 87.6 (85.2, 90.0) |
| 13 to 17 years | 90.1  (88.4, 91.7) | 89.2  (87.5, 91.0) | 91.2  (89.8, 92.6) | 88.8 (86.9, 90.8) | 86.0 (83.4, 88.6) | 85.3 (83.1, 87.6) | 82.0 (79.4, 84.7) | 82.6 (79.6, 85.6) | 84.6 (81.7, 87.4) | 85.9 (82.9, 89.0) |
| Hispanic | 89.9 (88.3, 91.5) | 89.7 (88.3, 91.1) | 91.8 (90.4, 93.2) | 90.3 (88.4, 92.3) | 82.6 (79.7, 85.6) | 86.1 (83.6, 88.6) | 82.5 (79.5, 85.5) | 85.5 (81.8, 89.3) | 83.2 (80.1, 86.3) | 79.7 (75.3, 84.1) |
| White | 95.0 (93.6, 96.5) | 94.1 (92.5, 95.6) | 95.6 (94.7, 96.6) | 93.5 (91.7, 95.2) | 92.5 (90.7, 94.3) | 92.2 (90.3, 94.2) | 89.7 (87.6, 91.8) | 91.8 (89.5, 94.1) | 89.9 (87.3, 92.5) | 91.6 (89.2, 94.1) |
| Black | 89.9 (87.3, 92.5) | 89.8 (87.3, 92.3) | 88.2 (84.9, 91.4) | 85.8 (81.9, 89.7) | 89.6 (86.5, 92.7) | 87.0 (83.1, 91.0) | 81.3 (75.7, 86.8) | 78.9 (72.7, 85.2) | 80.6 (74.4, 86.9) | 83.1 (76.0, 90.1) |
| Others | 93.3 (90.9, 95.8) | 90.6 (87.9, 93.4) | 90.5 (87.7, 93.3) | 87.7 (84.0, 91.3) | 89.3 (85.7, 92.9) | 90.1 (86.4, 93.8) | 87.3 (82.5, 92.1) | 85.7 (80.2, 91.2) | 83.4 (77.5, 89.4) | 91.1 (87.3, 94.8) |
| Private insurance | 94.7  (93.7, 95.7) | 93.6  (92.3, 94.9) | 94.4  (93.4, 95.4) | 92.4 (90.7, 94.1) | 91.1 (89.1, 93.0) | 91.8 (90.2, 93.5) | 87.8 (85.7, 89.8) | 90.7 (88.4, 93.0) | 89.2 (87.0, 91.4) | 91.1 (89.0, 93.3) |
| Public insurance | 91.4  (89.8, 92.9) | 90.7  (89.0, 92.3) | 92.1  (90.6, 93.6) | 90.3 (88.5, 92.1) | 87.5 (85.3, 89.6) | 88.1 (85.6, 90.6) | 85.9 (83.3, 88.6) | 84.6 (81.0, 88.3) | 83.2 (79.9, 86.6) | 83.1 (79.6, 86.7) |
| Uninsured | 78.4  (70.7, 86.0) | 79.4  (71.6, 87.2) | 72.9 (63.7, 82.1) | 66.4 (56.0, 76.8) | 68.2 (56.0, 80.4) | 65.3 (50.4, 80.2) | 59.9 (45.2, 74.6) | 48.7 (36.4, 60.9) | 54.8 (38.1, 71.5) | 53.6 (39.6, 67.6) |

**Supplement table 4. Distributions of grouped reasons for not having usual source of care of U.S. children 2015-2024: cells are percentage (95% CI).**

| Grouped  reasons | 2015 | 2016 | 2017 | 2018 | 2019 | 2020 | 2021 | 2022 | 2023 | 2024 |
| --- | --- | --- | --- | --- | --- | --- | --- | --- | --- | --- |
| Low perceived  Need | 47.9  (41.5, 54.3) | 49.7  (43.2, 56.3) | 53.3  (46.3, 60.2) | 49.7  (42.7, 56.7) | 55.4  (49.0, 61.7) | 49.2  (42.0, 56.3) | 58.1  (51.7, 64.5) | 57.4  (50.0, 64.7) | 60.9  (53.2, 68.5) | 54.1  (46.9, 61.3) |
| Preference or  self-management | 6.7  (3.8, 9.6) | 6.8  (4.0, 9.6) | 6.8  (3.5, 10.1) | 8.7  (4.9, 12.5) | 10.6  (7.1, 14.2) | 5.7  (2.5, 9.0) | 6.9  (3.5, 10.4) | 7.5  (3.2, 11.9) | 6.8  (3.4, 10.2) | 7.2  (2.6, 11.8) |
| Access or  continuity barrier | 39.3  (32.9, 45.7) | 37.0  (30.5, 43.5) | 31.5  (25.2, 37.8) | 34.1  (27.3, 40.9) | 24.4  (18.4, 30.4) | 32.1  (25.5, 38.8) | 26.9  (20.4, 33.4) | 22.5  (15.5, 29.5) | 21.8  (16.1, 27.5) | 25.0  (18.0, 31.9) |
| Other reasons | 6.1  (3.8, 8.4) | 6.5  (3.3, 9.7) | 8.5  (4.8, 12.1) | 7.5  (4.3, 10.8) | 9.7  (5.9, 13.4) | 13.0  (8.7, 17.2) | 8.0  (4.9, 11.2) | 12.6  (7.6, 17.6) | 10.6  (5.2, 15.9) | 13.8  (9.0, 18.5) |
